# Optimal Delivery Management for the Prevention of Early Neonatal SARS-CoV-2 Infection: Systematic review and Meta-analysis

**DOI:** 10.1101/2023.02.14.23285921

**Authors:** Christina S Chan, Juin Yee Kong, Rehena Sultana, Vatsala Mundra, Kikelomo L Babata, Kelly Mazzarella, Emily H Adhikari, Kee Thai Yeo, Jean-Michel Hascoët, Luc P Brion

## Abstract

**Objective:** Review how specific delivery management interventions (DMI) are associated with early neonatal SARS-CoV-2 infection (ENI) and neonatal death <28 days of life (ND).

**Design:** Systematic review and meta-analysis of individual patient-specific data from articles published 1 January 2020 - 31 December 2021 from Cochrane review databases, Medline and Google Scholar.

**Setting:** International publications specifying DMI, ENI, and ND.

**Patients:** Pregnant women infected with SARS-CoV-2 and their infants

**Main outcome measures:** Article inclusion criteria: 1) mothers with SARS-CoV-2 PCR positive status within 10 days before delivery or symptomatic at delivery with a positive test within 48 hours after delivery, 2) delivery method described, 3) infant SARS-CoV-2 PCR result reported. Primary outcomes were 1) ENI confirmed by positive neonatal PCR and 2) ND.

**Results:** Among 11,075 screened publications, 117 publications containing data for 244 infants and 230 mothers were included. Maternal and infant characteristics were pooled using DerSimonian-Laird inverse variance method. Primary outcome analyses were completed using logit transformation and random effect. Heterogeneity of included studies was evaluated with I^2^ statistics.

No routine care was described so comparison of DMI combinations to routine care was not possible. Sample size for each combination was too small to conduct any valid comparison of different DMI combinations.

**Conclusion:** Support for specific DMI in SARS-CoV-2 infected mothers is lacking. This review highlights the need for rigorous and multinational studies on the guidelines best suited to prevent transmission from mother to neonate.

**KEY MESSAGES:** *What is already known on this topic:* Several specific delivery management interventions (DMI) have been recommended for women with active SARS-CoV-2 to prevent early neonatal SARS-CoV-2 infection.

*What this study adds:* This systematic review shows that support for specific DMI in SARS-CoV-2 infected mothers is lacking.

*How this study might affect research, practice or policy:* This review highlights the need for rigorous and multinational studies on the guidelines best suited to prevent transmission from mother to neonate.

## SETTING

The first case of coronavirus disease of 2019 (COVID-19) was reported from China in November 2019. It was later found to be caused by a novel virus named severe acute respiratory syndrome coronavirus 2 (SARS-CoV-2) ^1, 2^. On 30 January 2020, the World Health Organization ^2^ declared the outbreak a Public Health Emergency of International Concern ^3^. By 11 March 2020, COVID-19 was characterized as a global pandemic ^2^. The rapid progression of COVID-19/SARS-CoV-2 dramatically impacted healthcare, especially for pregnant women and infants at increased risk for maternal death, severe maternal morbidities, and neonatal morbidities^4^. With significant numbers of pregnant women infected with SARS-CoV-2, lack of consistent vaccine availability in many countries, and vaccine hesitancy especially for pregnant women, evidence for practice recommendations designed to reduce transmission to infants remains a pressing concern.

The mode and significance of SARS-CoV-2 transmission from mother to fetus or infant remains unclear. Confirmation of diagnosis and source of transmission is challenging, given the subtle presentation of symptoms and clinical course in newborn infants and the possible close contact of multiple caregivers who may be infectious ^5^. With available case reports, it has been challenging to accurately determine exact timing or route of transmission as this would require extensive testing including immunocytochemistry, in situ hybridization and/or RT-PCR in all possible pregnancy and delivery material (placenta, amniotic fluid, vaginal secretions, colostrum, blood, stools, urine, nasopharyngeal secretions in the mother), and serially timed PCR sampling of the infant and sampling among all caregivers.

## PATIENTS

To date, there has been no systematic review or study comparing combinations of delivery management interventions (DMI) with routine care and the risk of early neonatal infection (ENI) or neonatal death before 28 days of life (ND). Differing opinions from various expert consensus guidelines make the best choices for DMI challenging. Many suggest shared decision-making between healthcare providers and patients for management during the perinatal period ^6^. With the ubiquitous burden of this disease, it remains important to assess DMI that could minimize ENI and ND while ensuring safety and optimal care to both mother and infant.

### IN UTERO INFECTION

In utero infections have now been documented. Between 1% and 15% of adults with COVID-19 have RNAaemia and pregnant women are more susceptible to COVID-19 than the general population^1, 7^. Physiologic mechanisms for in utero infections include angiotensin-converting enzyme 2 (ACE2), the receptor of SARS-CoV-2 as an integral component of human to human transmission that is ^8^ expressed in maternal-fetal interface cells ^9^, placenta ^10, 11^ as well as several fetal organ cells ^9, 12^. Detection of SARS-CoV-2 RNA in pharyngeal or stool specimens of infants born to mothers with a remote history of SARS-CoV-2 infection during pregnancy, suggests possible congenital infection as persistence of the virus is expected for up to five weeks after onset of symptoms ^13^. In-utero infection with SARS-CoV-2 has been suggested via the amniotic fluid ^14^ as well as placenta ^15^. The detection of IgM in infants shortly after birth despite failure to isolate viral RNA also suggests the transmission of infection prior to delivery ^16–19^.

### PERINATAL INFECTION

In addition, perinatal transmission of SARS-CoV-2 may also be possible during labor and delivery. In the immediate postnatal period, direct exposure of infants to infected maternal stool, blood or amniotic fluid can increase transmission risk to the infant ^20^. There is also theoretical risk of airborne transmission from mothers who have experienced a recent aerosol-generating procedure such as intubation or electrocautery of surgical wound ^21^. There has also been a suggestion that the second stage of labor, while not typically described as an aerosol-generating event, is a period when mothers may be breathing heavily, shouting, coughing, or vomiting - all activities that increase risk of aerosol exposure ^20^.

### EARLY NEONATAL INFECTION

ENI may be evident between 12 hours of life and 15 days of life with the average incubation period of five days ^22^. For this review’s purposes, we define ENI as neonatal SARS-CoV-2 PCR-positive between 12 hours and 10 days of life. This may include in utero and perinatal infection. Beyond this period, delivery management is less likely to affect whether an infant is SARS-CoV-2-positive.

## INTERVENTIONS

### EVOLVING GUIDELINES AND PRACTICES

In addition to the important consideration of ENI and ND, infection control practices recommended by international, national, regional and hospital-specific guidelines have had unintended consequences. For example, interventions intended to prevent maternal-neonatal transmission of virus have changed the routine practice of a planned normal birth. The decision for an unplanned caesarean section delivery, separation of infants right after birth, strict isolation and visitation policies have resulted in a variety of medical and psychosocial effects on the mother-infant dyad and family.

Different countries, international/national/regional organizations and hospitals have published recommendations on the management of a SARS-CoV-2-positive mother during labor, delivery, and for her infant (Figure 1). All recommendations were designed to balance alterations in delivery management to reduce SARS-CoV-2 transmission to the infant, with routine care recommendations that promote bonding and are beneficial to mother and infant. In the absence of data, guidelines were created ad hoc and with significant variation in practice ^23^. Several of these recommendations for delivery of mothers with active SARS-CoV-2 were based on other infectious diseases (e.g. influenza), with little knowledge about the incidence and clinical presentation of neonatal SARS-CoV-2 infection.

**Figure 1.**
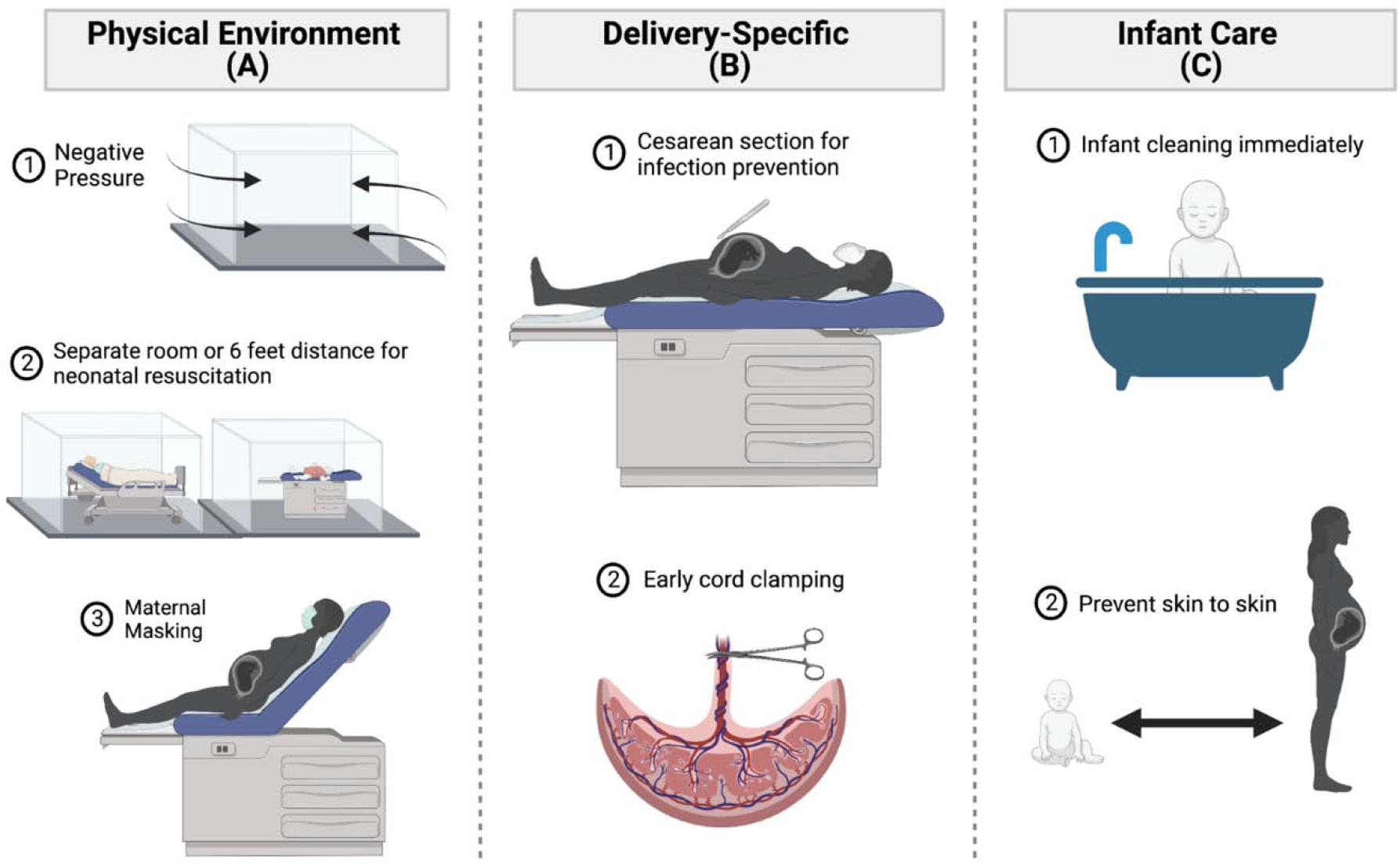
Depiction of delivery management interventions (DMI). At minimum a single intervention from category A – Physical Environment, category B – Delivery-Specific, or category C – Infant Care in combination or alone are evaluated for the primary outcomes and are compared to routine care. The different categories of interventions are 1) Combo A alone, 2) Combo B alone, 3) Combo C alone, 4) Combo A + B + C, 5) Combo A + B not C, 6) Combo B + C not A, and 7) Combo A + C not B.

## OBJECTIVE

We intended to describe the frequency of ENI and ND, and their association with different DMI combinations as suggested by country or organization-specific guidelines, when compared with routine delivery practice.

We categorized DMI into three broad categories: 1) physical environment, 2) delivery-specific, and 3) infant-care practices (Figure 1). Within each category there are individual interventions intended for infection prevention at various times during delivery. In different countries and birth centers, these interventions were applied in different combinations because of unique circumstances dictating resource allocation, disease burden, and local practice. We have described these combinations with reference to several country-specific guidelines that were available at the inception of this review (see Appendix 1) ^24^.

### FACTORS NOT ADDRESSED

Enteral feeding practices and post-partum behaviors also likely impact ENI and ND. As these are detailed and nuanced decisions often made independent of the DMI, enteral feeding are beyond the scope of this review.

It was also difficult to completely remove the risk of a SARS-CoV-2 positive healthcare worker or family member for ENI and ND. For many countries, the possibility of a healthcare worker contributing to early infection is a highly litigious aspect of infant care and will not be reported in case reports or databases. For pragmatic reasons, we will not be addressing potential transmission of SARS-CoV-2 from family members or healthcare workers to the neonate.

## DESIGN

The protocol for this review was submitted to PROSPERO ID CRD42021267892. Articles published 1 January 2020 - 31 December 2021 from Cochrane review databases, Medline and Google Scholar were searched. Primary screening was conducted by 2 reviewers (CSC and JYK) using specific search criteria (Appendix 2).

We included the following studies (listed in the order of the strength of evidence):

1. Randomized controlled trials
2. Quasi-randomized trials
3. Cohort studies
4. Case-control studies
5. Cross-sectional studies
6. Case series/case reports

Specific inclusion criteria included

1. mothers with SARS-CoV-2 PCR positive status within 10 days before delivery or symptomatic at delivery with a positive test within 48h after delivery
2. delivery method described
3. infant SARS-CoV-2 PCR result known

## MAIN OUTCOME MEASURES

Types of DMI (see Appendix 3)

A. Physical environment (aerosolization and droplet management):
  1. Negative pressure in delivery area
  2. Separate room from delivery room
  3. Distance of ≥ 6 feet from mother during resuscitation
  4. Maternal masking during delivery compared to no maternal masking during delivery
B. Delivery-specific interventions (minimization of contact during delivery with maternal fluids):
  1. Caesarean section for infection prevention
  2. Early cord clamping compared to delayed cord clamping
C. Infant care practices (minimizing infant skin contact):
  1. Infant cleaning/decontamination as soon as possible after resuscitation compared to routine skin care
  2. Prevention of skin-to-skin contact after stabilization compared to skin-to-skin contact after stabilization

Primary outcomes

1. Confirmation of early neonatal SARS-CoV-2 infection by a positive PCR on any neonatal samples taken at 12 hours and up to 10 days of life.
2. Neonatal death occurring before 28 days of life.

Secondary outcomes were selected by their potential relationship with SARS-CoV-2 infection:

1. Maternal characteristics:
  a. Age
  b. maternal symptom severity (Appendix 4)
    i. Asymptomatic
    ii. Mild
    iii. Moderate
    iv. Severe
    v. Critical
2. Delivery metrics:
  a. Gestational age
  b. Infant sex
  c. Birth weight
  d. Apgar scores at one minute and five minutes
3. Clinical status in SARS-CoV-2 positive infants (Appendix 4)
  a. Asymptomatic
  b. Mild
  c. Moderate
  d. Severe
  e. Critical

## DATA COLLECTION AND ANALYSIS

Cochrane search was used for the initial search (initial strategy). A combination of Medline and Google Search (updated strategy) was used by two review authors for later search and screening (JMH, LPB). Comparison of the initial and updated strategies found 96.5% overlap of the two strategies for publication identification and inclusion. Two review authors (CSC, JYK) independently screened all titles, abstracts and full-text reports for eligibility. They resolved any differences in decision between authors by discussion leading to consensus. They recorded the selection process in sufficient detail to complete a PRISMA flow diagram (Figure 2). They extracted data using a pilot-tested Microsoft Excel spreadsheet and a comprehensive list of included articles can be found in Appendix 5.

**Figure 2.**
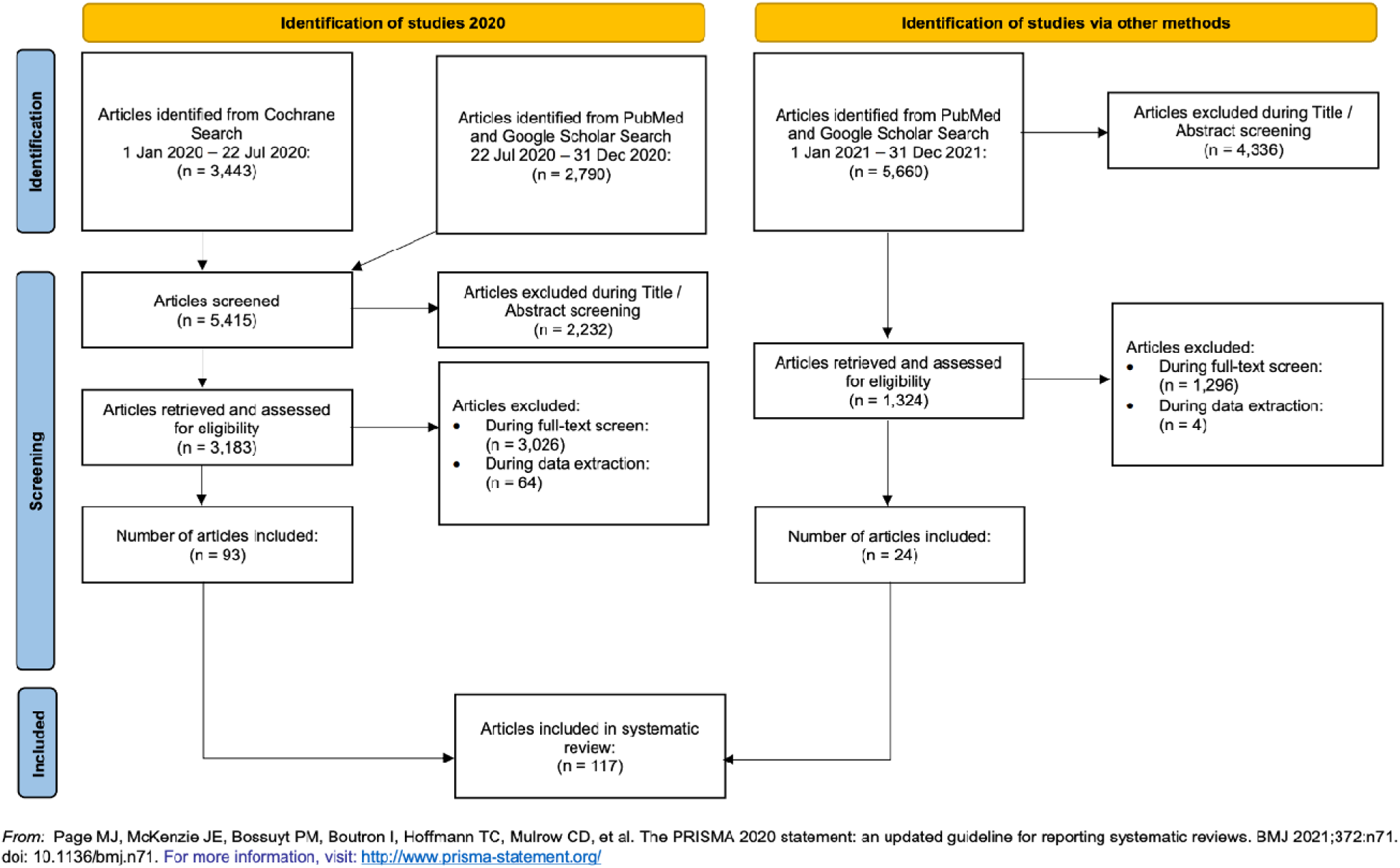
PRISMA Flow Diagram of included studies. Search strategies included Cochrane search 1 January 2020 to 22 July 2020, PubMed and Google Scholar Search 22 July 2020 to 31 December 2020, and PubMed and Google Scholar Search 1 January 2021 to 31 December 2021.

Primary outcomes were ENI and ND. Both outcomes were treated as binary variables. All analyses were carried out using individual patient level data. All maternal and infant characteristics were pooled using DerSimonian-Laird inverse variance method. Pooled results were expressed as percentages with 95% confidence interval (95%CI) for categorical variables and as mean (95%CI) or median (95%CI), whichever appropriate, for continuous variables. Primary outcome analyses were carried out using logit transformation and random effect. Continuity correction was applied for all pooled results if any included study has 0 event. The heterogeneity of included studies was evaluated with I^2^ statistics, considering a value less than 25% as low heterogeneity, 50-75% as medium heterogeneity and greater than 75% were considered as high heterogeneity. All the analyses were carried out using SAS version 9.4 (SAS Institute; Cary, NC, USA) and R (‘meta’ library).

## RESULTS

Of 11,075 publications initially screened, 117 articles were selected for inclusion (Figure 2). Data used for this analysis from the 117 publications included 243 infants and 231 mothers.

Clinical characteristics are described in Table 1. The mean maternal age was 31.2 years. The majority, 31.3% [29.97, 32.49], of these women infected with SARS-CoV-2 showed moderate illness severity. 79% [69.2, 88.9] of women experienced cesarean-section delivery. The mean gestational age was 35.6 [34.2, 37.1] weeks. 58% [44.6, 72.2] of these cases were male infants.

**Table 1.**
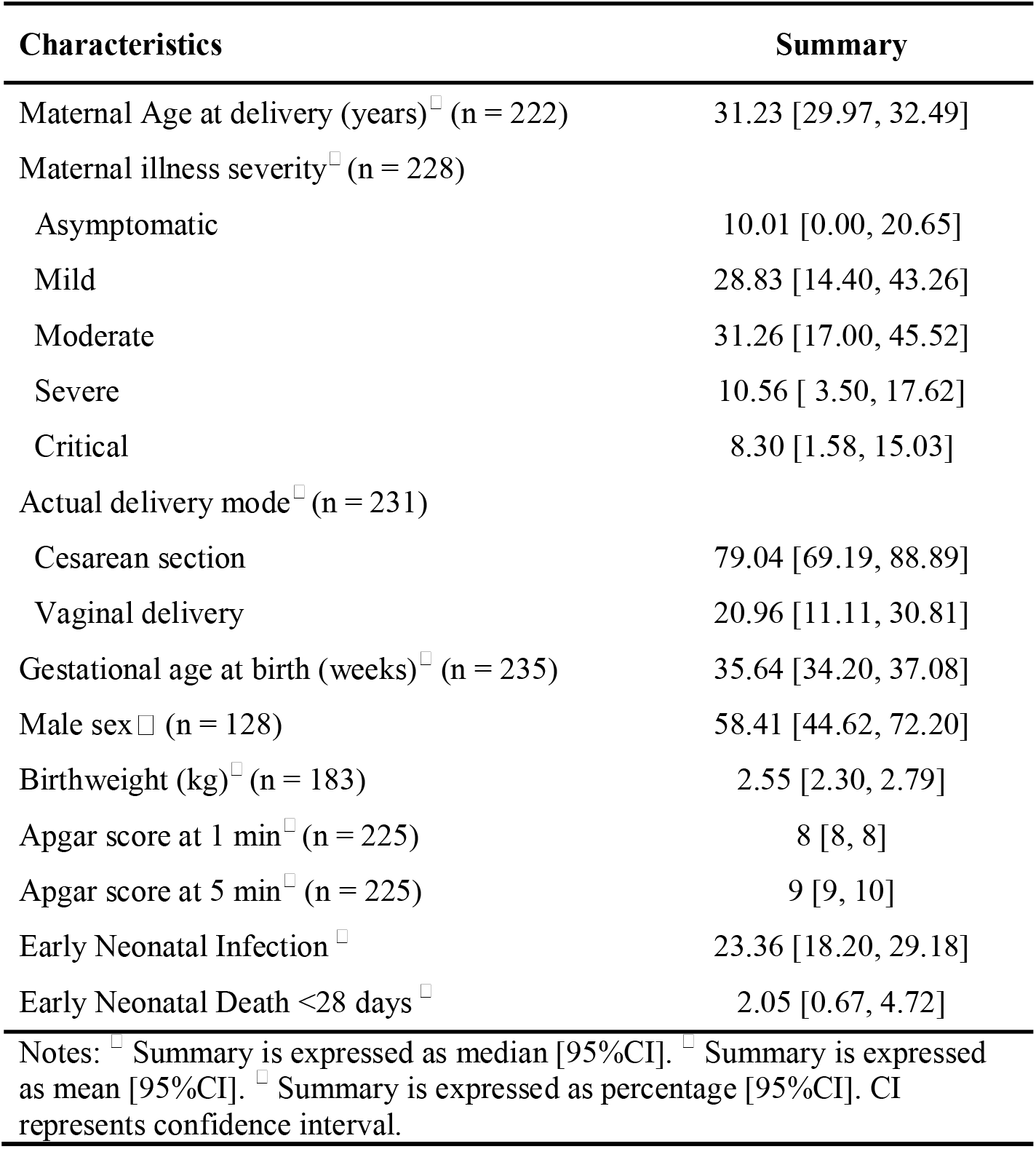
Clinical characteristics of maternal-infant pairs affected by perinatal COVID-19.

| Characteristics | Summary |
| --- | --- |
| Maternal Age at delivery (years) <sup>□</sup> (n = 222) | 31.23 [29.97, 32.49] |
| Maternal illness severity <sup>□</sup> (n = 228) |  |
| Asymptomatic | 10.01 [0.00, 20.65] |
| Mild | 28.83 [14.40, 43.26] |
| Moderate | 31.26 [17.00, 45.52] |
| Severe | 10.56 [ 3.50, 17.62] |
| Critical | 8.30 [1.58, 15.03] |
| Actual delivery mode <sup>□</sup> (n = 231) |  |
| Cesarean section | 79.04 [69.19, 88.89] |
| Vaginal delivery | 20.96 [11.11, 30.81] |
| Gestational age at birth (weeks) <sup>□</sup> (n = 235) | 35.64 [34.20, 37.08] |
| Male sex <sup>□</sup> (n = 128) | 58.41 [44.62, 72.20] |
| Birthweight (kg) <sup>□</sup> (n = 183) | 2.55 [2.30, 2.79] |
| Apgar score at 1 min <sup>□</sup> (n = 225) | 8 [8, 8] |
| Apgar score at 5 min <sup>□</sup> (n = 225) | 9 [9, 10] |
| Early Neonatal Infection <sup>□</sup> | 23.36 [18.20, 29.18] |
| Early Neonatal Death <28 days <sup>□</sup> | 2.05 [0.67, 4.72] |
| Notes: <sup>□</sup> Summary is expressed as median [95%CI]. <sup>□</sup> Summary is expressed as mean [95%CI]. <sup>□</sup> Summary is expressed as percentage [95%CI]. CI represents confidence interval. |  |

The mean birthweight was 2550 grams [2300, 2790]. Median Apgar scores were 8 [8, 8] at 1 minute, and 9 [9, 10] at 5 minutes. ENI was described in 23.4% [18.2, 29.18] of cases. Early neonatal death <28 days was reported in 2.1% [0.67, 4.72] of cases.

Total cases of each delivery management intervention are depicted in Table 2

**Table 2.** Totals of infants and mothers with described delivery management interventions.

| **Delivery Management Intervention** | **Number of infants or mothers** | **Percentages (95% CI)** |
| --- | --- | --- |
| **A: Physical environment (aerosolization and droplet management)** | **67** |  |
| Negative pressure in delivery room | 21/25 | 85.47 [73.15, 97.79] |
| Separate room for resuscitation of infant or Distance of $\geq 6$ feet from mother during resuscitation | 32/34 | 94.33 [84.23, 100.00] |
| Maternal masking during delivery | 38/38 | 100.00 [90.39, 100.00] |
| **B: Delivery-specific interventions (minimization of contact during delivery with maternal fluids)** | **59** |  |
| Caesarean section for infection prevention purpose | 41/58 | 47.27 [28.72, 65.83] |
| Early cord clamping | 18/18 | 100.00 [86.03, 100.00] |
| **C: Infant care practices (minimization of infant skin contact)** | **79** |  |
| Infant cleaning or decontamination right after resuscitation | 4/5 | 80.00 [40.80, 100.00] |
| Prevent skin-to-skin after stabilization | 79/79 | 100.00 [93.88, 100.00] |
| Notes:<br>• Denominator represents # of participants with reported (yes/no) particular exposures while numerator represents number of mothers /children had that particular exposure. Participants were not included in the analysis, if there was any missing exposure.<br>• DerSimonian-Laird inverse variance estimator were used to pool the results. Simple approximation with continuity correction of 0.5 in studies with zero cell frequencies was applied. Variables were expressed as percentages (95%CI). CI represents confidence interval. |  |  |

67 cases reported on the physical environment designed to minimize aerosolization and droplets from the infected mother. Among 25 cases with information on the use of negative pressure in the delivery room, this DMI was used in 21 (85.5% [73.15, 97.79]). Among 34 cases with information on the use of a separate room for infant resuscitation or a distance of > 6 feet from the mother during the resuscitation, this DMI was used in 32 (94.3% [84.23, 100]). 38 cases contained information about maternal masking during delivery and all reported use of this DMI (100% [90.39, 100]).

Among 59 cases with information on delivery-specific interventions designed to minimize contact during delivery with maternal secretions, use of cesarean section for infection prevention purposes was reported in 58: 41 reported use of this DMI (47.3% [28.72, 65.83]). 18 cases reported on early cord clamping and all reported use of this DMI (100% [86.03, 100]).

Among 79 cases with information on infant care practices designed to minimize infant skin contact with infected secretions, 5 cases reported specifically on infant cleaning or decontamination right after the resuscitation: 4 reported use of this DMI (80% [40.8, 100]). Among 79 cases with information on preventing skin-to-skin with the mother after infant stabilization, all reported use of this DMI (100% [93.88, 100]).

Primary outcomes are described in Table 3. Combination A describes physical environmental changes, combination B describes delivery-specific interventions and combination C describes infant care interventions. Combination 1) A only, 2) B only, 3) C only, 4) A plus B plus C, 5) A plus B without C, 6) B plus C without A, and 7) A plus C without B were evaluated for incidence of the primary outcomes ENI and ND compared to routine care. Pooled frequencies of ENI and ND were reported as random effect with all DMI alone and DMI combinations (7 total comparisons). No routine care was described in these publications so the primary comparison of DMI combinations to routine care was not possible. Sample size for each combination was too small to conduct any valid comparison of the 6 different DMI combinations.

**Table 3.**
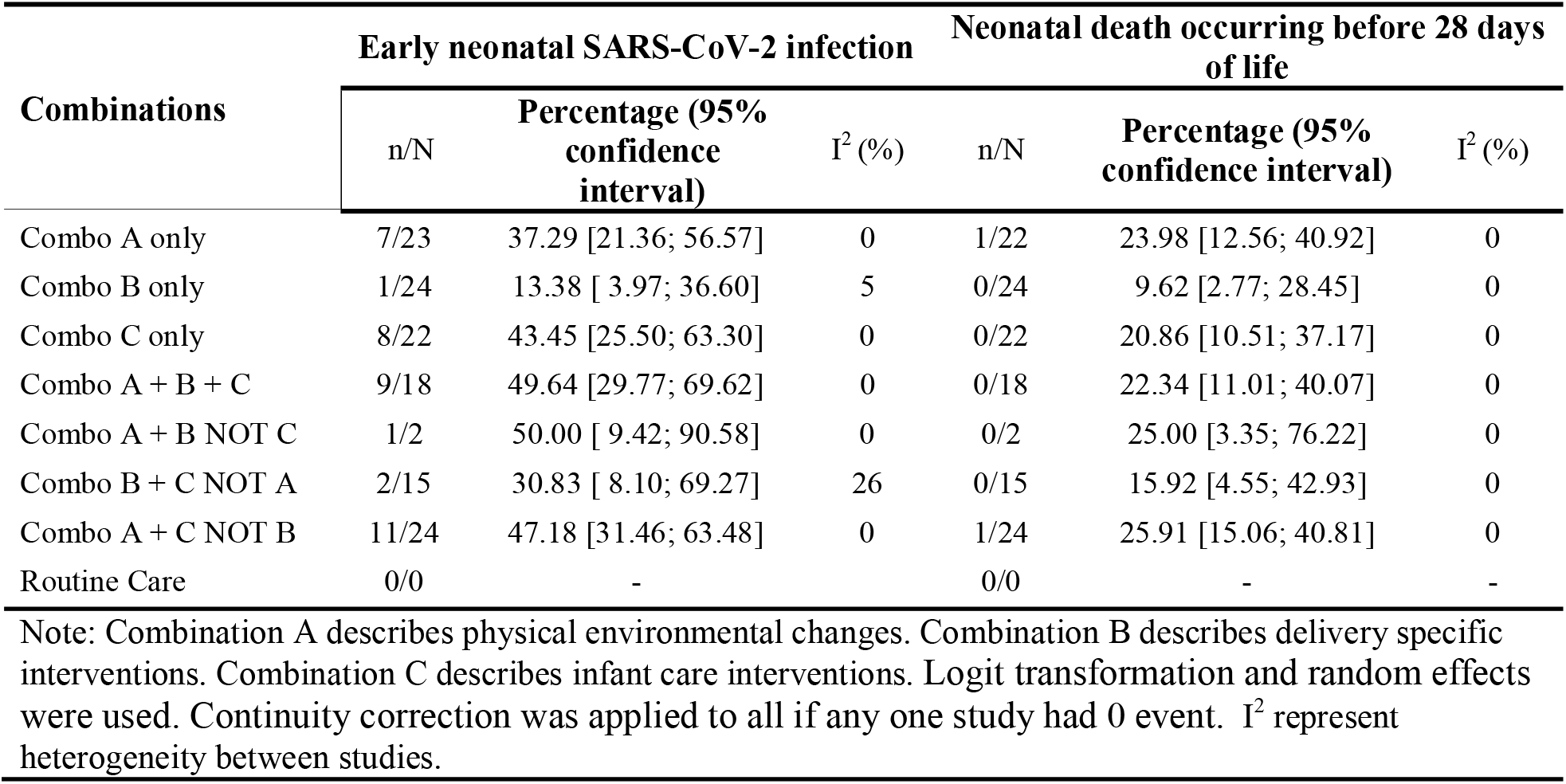
Primary outcomes of early neonatal infection and neonatal death by delivery management interventions *individually by category and in combinations of different categories*.

| Combinations | Early neonatal SARS-CoV-2 infection |  |  | Neonatal death occurring before 28 days of life |  |  |
| --- | --- | --- | --- | --- | --- | --- |
|  | n/N | Percentage (95% confidence interval) | I<sup>2</sup> (%) | n/N | Percentage (95% confidence interval) | I<sup>2</sup> (%) |
| Combo A only | 7/23 | 37.29 [21.36; 56.57] | 0 | 1/22 | 23.98 [12.56; 40.92] | 0 |
| Combo B only | 1/24 | 13.38 [ 3.97; 36.60] | 5 | 0/24 | 9.62 [2.77; 28.45] | 0 |
| Combo C only | 8/22 | 43.45 [25.50; 63.30] | 0 | 0/22 | 20.86 [10.51; 37.17] | 0 |
| Combo A + B + C | 9/18 | 49.64 [29.77; 69.62] | 0 | 0/18 | 22.34 [11.01; 40.07] | 0 |
| Combo A + B NOT C | 1/2 | 50.00 [ 9.42; 90.58] | 0 | 0/2 | 25.00 [3.35; 76.22] | 0 |
| Combo B + C NOT A | 2/15 | 30.83 [ 8.10; 69.27] | 26 | 0/15 | 15.92 [4.55; 42.93] | 0 |
| Combo A + C NOT B | 11/24 | 47.18 [31.46; 63.48] | 0 | 1/24 | 25.91 [15.06; 40.81] | 0 |
| Routine Care | 0/0 | - |  | 0/0 | - | - |

Note: Combination A describes physical environmental changes. Combination B describes delivery specific interventions. Combination C describes infant care interventions. Logit transformation and random effects were used. Continuity correction was applied to all if any one study had 0 event. I<sup>2</sup> represent heterogeneity between studies.

Subgroup analyses of maternal illness severity and gestational age effects on ENI and ND are described in Appendix 6. Neither maternal illness severity nor gestational age had statistically significant effects on ENI or ND.

## DISCUSSION

This is the first systematic review of published cases describing specific interventions in delivery management that were recommended during the pandemic to reduce the possibility of ENI and ND before 28 days of life. While the pandemic has changed with the introduction of several vaccines and the exposure of most worldwide populations to SARS-CoV-2, the question of how best to protect infants from an airborne infection remains relevant.

Our review highlights significant publication biases inherent during the active phases of the pandemic including preferential publication of deliveries where management was altered without similar descriptions of outcomes for deliveries conducted in routine fashion, preferential publication of cases involving ENI and ND, and publication of multiple reports that did not include specific information regarding routine delivery management practices. These biases in publication along with limitations including lack of randomization, limited case numbers, and limited data on postpartum care contribute to our ongoing inability to provide evidence for best practice delivery management modifications for reducing ENI and ND.

Given our findings, it is uncertain what contribution delivery management interventions may play into neonatal outcomes. Instead, ENI and ND may be influenced by maternal illness severity prior to delivery, viral load during the pregnancy, and post-partum practices where mild to moderately symptomatic mothers could cohort with their infants.

## CONCLUSIONS

Comparison of DMI to routine care was not possible in this systematic review. More data is needed to accurately assess the effect of these DMI on ENI and ND. We recommend investing in large, multinational, prospective database creation with specific attention to DMI to more systematically address the question of optimal delivery management for prevention of airborne transmission from mother to infant.

## Data Availability

All data produced in the present study are available upon reasonable request to the authors

## Abbreviations

DMI: delivery management interventions
ENI: early neonatal SARS-CoV-2 infection
ND: neonatal death (<28 days of life)
SARS-CoV-2: severe acute respiratory syndrome coronavirus 2
COVID-19: coronavirus disease of 2019, the disease caused by SARS-CoV-2 virus

## Acknowledgements

There was no funding for this publication.

## Contributions of authors

CSC and JYK wrote the first version of the protocol and subsequent manuscript drafts. RS provided statistical support and contributed to methods. VM and KM contributed to data extraction. EHA provided obstetric expertise. JMH and LPB reviewed and extracted articles using the updated search strategy. All other authors have contributed to and approved the current version.

## Sources of support

### Internal sources

UT Southwestern Medical Center, Dallas, TX, USA

Division of Neonatal-Perinatal Medicine

### External sources

Vermont Oxford Network, USA

Cochrane Neonatal Reviews are produced with support from Vermont Oxford Network, a worldwide collaboration of health professionals dedicated to providing evidence-based care of the highest quality for newborn infants and their families.

