## Supplementary material for "Optimal Delivery Management for the Prevention of Early Neonatal SARS-CoV-2 Infection: Systematic review and Meta-analysis": Data Supplement

**SUPPLEMENTAL MATERIAL**

**Appendix 1. Country specific guidelines for reducing SARS-CoV2 exposure.**

Delivery management alterations described by guidelines. This is not a comprehensive review of all guidelines regarding obstetric and neonatal practice. This is a pragmatic review for select recommendations for or against practices that have been described in case reports regarding management of SARS-CoV-2-positive mothers and their infants designed to reduce transmission to the newborn.

| **Delivery Management Alternation** | **Guidelines Recommending Practice** | **Guidelines Recommending Against Practice** |
| --- | --- | --- |
| Negative pressure in delivery room or resuscitation room | AAP 2020;  China 2020a;  China 2020b;  China 2020c;  China 2020g;  France 2020a;  France 2020b;  France 2020c;  SOAP SMGM 2020; |  |
| Separate room from delivery room for neonatal resuscitation or distance from mother during resuscitation of six feet or more | AAP 2020;  China 2020d;  China 2020f;  China 2020g;  CPS 2020;  France 2020a;  France 2020c;  India 2020;  Saudi 2020; |  |
| Mother masked during labor/delivery | AAP 2020;  China 2020d;  France 2020a;  France 2020c;  RCOG 2020;  SOGC 2020;  Spain 2020; |  |
| Cesarean delivery for infection prevention purposes |  | AAP 2020;  ACOG 2020;  CDC 2020;  China 2020i;  France 2020a;  France 2020c;  India 2020;  RCOG 2020;  Saudi 2020;  SOGC 2020; |
| Early cord clamping | China 2020a;  China 2020b;  China 2020c;  China 2020d;  China 2020f;  China 2020g;  Saudi 2020;  Spain 2020 | AAP 2020;  ACOG 2020;  CDC 2020;  CPS 2020;  India 2020;  RCOG 2020;  SOGC 2020; |
| Infant cleaning or decontamination as soon as possible after resuscitation | None documented | None documented |
| Avoidance of skin-to-skin contact | None documented | None documented |

1. AAP 2020: American Academy of Pediatrics. Management of infants born to mothers with COVID-19. downloads.aap.org/AAP/PDF/COVID%2019%20Initial%20Newborn%20Guidance.pdf (accessed prior to 2 April 2020).
2. ACOG 2020: The American College of Obstetricians and Gynecologists. COVID-19 FAQs for obstetricians-gynecologists, obstetrics. www.acog.org/clinical-information/physician-faqs/covid-19-faqs-for-ob-gyns-obstetrics (accessed 23 March 2020).
3. China 2020a: Kaixu W, Yu H, Fang L, Yuan S, Children’s Hospital of Chongqing Medical University, National Clinical Research Center for Child Health and Disorders, International Science and Technology Cooperation Base of Child Development and Critical Disorders, Ministry of Education Key Laboratory of Child Development and Disorders, Key Laboratory of Child Infection and Immunity, Chongqing. Epidemiological characteristics of corona virus disease 2019 and SARS and Prevention and Control of Corona Virus Disease 2019 in Newborns [新型冠状病毒肺炎与严重急性呼吸综合征流行特征及新生儿新型冠状病毒肺炎防控策略]. Journal of Pediatric Pharmacology 2020;26(4):46-48. [doi: 10. 13407 /j. cnki. jpp. 1672-108X. 2020. 04. 015]
4. China 2020b: Fang L, Yuan S, Department of Neonatology, Children’s Hospital of Chongqing Medical University/Ministry of Education Key Laboratory of Child Development and Disorders/National Clinical Research Center for Child Health and Disorders/China International Science and Technology Cooperation Base of Child Development and Critical Disorders. Summary of consensus documents: prevention and control of 2019 novel coronavirus infection in newborn infants [新生儿 2019 新型冠状病毒感染防控共识方案要点解析]. ChongQing Medicine 2020. [kns.cnki.net/kcms/detail/50.1097.r.20200213.1614.011.html]
5. China 2020c: Yang Jie, Chen Yun Bin (杨杰，陈运彬), Guangdong Neonatal ICU Medical Quality Control Center (广东省新生儿ICU质量控制中心). 广东省新型冠状病毒感染疫情下新生儿科诊疗防控预案. Journal of Practical Medicine 2020. [kns.cnki.net/kcms/detail/44.1193.R.20200423.1322.002.html]
6. China 2020d: Health Commission of Guangxi Zhuang Autonomous Region, China. 确诊和疑似新冠肺炎孕产妇安全助产新生儿防治应急预案. wsjkw.gxzf.gov.cn/zwgk/zfxxgkml/wsjszh/fybj/2020/0219/1747.html (accessed 19 February 2020).
7. China 2020e: Zhou WH, Working Group for the Prevention and Control of Neonatal 2019-nCoV Infection in the Perinatal Period of the Editorial Committee of Chinese Journal of Contemporary Pediatrics. Perinatal and neonatal management plan for prevention and control of 2019 novel coronavirus infection (1st edition) [围产新生儿新型冠状病毒感染防控管理预案（第一版)]. www.zgddek.com/EN/abstract/abstract14940.shtml (accessed 30 January 2020).
8. China 2020f: Yue Shao Jie (corresponding author) Hunan Neonatal Medical Quality Control Center. The programme for prevention and control of SARS-CoV-2 infection in neonates in Hunan Provence [湖南省新生儿新型冠状病毒感染防治方案]. China Journal of Modern Medicine 2020;30(4):65-71. [DOI: 10.3969/j.issn.1005-8982.2020.04.012]
9. China 2020g: Chen M-Y, Lu X, Qian X, School of Public Health, Fudan University. An overview of official measuers for COVID-19 prevention and control in maternal and neonatal population in China [我国部分地区孕产妇和新生儿新型冠状病毒感染防控措施综述]. Shanghai Journal of Preventive Medicine 2020. [DOI：10.19428/j.cnki.sjpm.2020.20249] [kns.cnki.net/kcms/detail/31.1635.r.20200525.1929.003.html]
10. China 2020h: Li Yu Hong, CHen Qian et al (李玉红，陈前，张沛佩，万振霞，王庆玲，芦庆花，毛青青，唐敬海，黄磊), ShanDong Province Maternal and Child Health Centre (山东省妇幼保健院，济南). 新生儿新型冠状病毒感染防控策略. Shandong Medicine 2020;60(10):110-113. [DOI: 10.3969 /j.issn.1002-266X.2020.10.029]
11. China 2020i: Fong Ling (冯玲) (corresponding author), Wuhan Tongji Hospital (Tongji Medical College of Huazhong University of Science and Technology). Guidelines for management of pregnant women and neonates with novel coronavirus infection in Wuhan Tongji Hospital during the epidemic period [武汉同济医院-新型冠状病毒感染的肺炎流行期间孕产妇及新生儿管理指导意见(第一版)]. China Journal of Obstetrics Emergency 2020;9(1):1-4. [DOI: 10.3877/cmaj.issn.2095-3259.2020.01.001]
12. CPS 2020: Canadian Paediatric Society. Delivery room considerations for infants born to mothers with suspected or proven COVID-19. www.cps.ca/en/documents/position/delivery-room-considerations-infants-born-to-mothers-with-suspected-or-proven-covid-19 (accessed 6 May 2020).
13. France 2020a: Collège National des Gynécologues et Obstétriciens Français (CNGOF). Prise en charge aux urgences maternite d'une Patiente enceinte suspectee ou infectee par le coronavirus (COVID 19) (Management of a pregnant patient suspected or infected with coronavirus (COVID 19)). www.cngof.fr/coronavirus-go-cngof (accessed 17 March 2020).
14. France 2020b: Société Francaise d'Hygiène Hospitaliere. Avis relatif aux mesures d’hygiène pour la prise en charge d’un patient considéré comme cas suspect, possible ou confirmé d’infection à 2019-nCoV. www.sf2h.net/wp-content/uploads/2020/01/Avis-prise-en-charge-2019-nCo-28-01-2020.pdf (accessed 28 January 2020).
15. France 2020c: Académie Nationale de Médecine. Covid-19, pregnancy and childbirth. www.academie-medecine.fr/wp-content/uploads/2020/03/20.3.30-Communiqu%C3%A9-Covid19-et-grossesse-ENG.pdf (accessed 30 March 2020).
16. India 2020: Federation of Obstetric & Gynaecological Societies of India National Neonatology Forum, India Indian Academy of Pediatrics. Perinatal-neonatal management of COVID-19. www.nnfi.org/assests/upload/announcement-pdf/Summary_FOGSI_NNF_IAP_Perinatal_Covid19-ver2May11merged.pdf (accessed prior to 10 July 2020).
17. RCOG 2020: Department of Health and Social Care (DHSC), Public Health Wales (PHW), Public Health Agency (PHA) Northern Ireland, Health Protection Scotland (HPS), Public Health Scotland, Public Health England and NHS England (official guidance). COVID-19: infection prevention and control guidance. www.rcog.org.uk/en/guidelines-research-services/guidelines/coronavirus-pregnancy/ (accessed 27 April 2020).
18. Saudi 2020: Saudi MOH Guideline for neonate born to mothers with suspected or confirmed COVID-19 infection. www.moh.gov.sa/Ministry/MediaCenter/Publications/Documents/Guideline-for-Neonate-born-to-COVID-19-Mother.pdf (accessed prior to 10 July 2020).
19. SOAP SMGM 2020: Society for Obstetric Anesthesia and Perinatology (SOAP), Society for Maternal-Fetal Medicine (SMFM). Labor and delivery COVID-19 considerations. soap.org/education/provider-education/expert-summaries/interim-considerations-for-obstetric-anesthesia-care-related-to-covid19/ (accessed prior to 14 April 2020).
20. SOGC 2020: Society of Obstetricians and Gynaecologists of Canada (SOGC). Updated SOGC committee opinion – COVID-19 in pregnancy. www.sogc.org/en/content/featured-news/Updated-SOGC-Committee-Opinion__COVID-19-in-Pregnancy.aspx (accessed 13 March 2020).
21. Spain 2020: Spanish Society of Neonatology. Recommendations for management of newborns for SARS-CoV-2 infection. www.seneo.es/images/site/noticias/home/Recomendaciones_SENeo_COVID-19_v6_eng.pdf (accessed prior to 10 July 2020).

**Appendix 2. Search strategies**

We accepted peer-reviewed published studies, both in full article and abstract forms as long as the minimum inclusion criteria are met: maternal SARS-CoV-2 PCR status known positive within 10 days of delivery or if symptomatic and status is known positive within 48h of delivery, delivery method, and infant SARS-CoV-2 PCR status known. We did not apply language restrictions. If non-English records were identified, we obtained a translation of the full-text report of the record to assess eligibility or had an author reviewer fluent in that language evaluate for eligibility.

**Initial Search Strategy**

We used the standard search strategy of Cochrane Neonatal to search Cochrane Central Register of Controlled Trials (CENTRAL 2020, Issue 7) in the Cochrane Library; Ovid MEDLINE(R) and Epub Ahead of Print, In-Process & Other Non-Indexed Citations, Daily and Versions(R); Cochrane COVID-19 Study Register; CINAHL; World Health Organization (WHO) COVID-19 Global literature on coronavirus disease; and the Centers for Disease Control and Prevention COVID-19 Research Articles Database on 23 July 2020. We also searched clinical trials databases and reference lists of retrieved articles for randomized controlled trials and quasi-randomized trials.

We conducted a comprehensive search on 23 July 2020 including:

1. Cochrane Central Register of Controlled Trials (CENTRAL 2020, Issue 7) in the Cochrane Library;
2. Cochrane COVID-19 Study Register, via CRS Web;
3. Ovid MEDLINE(R) and Epub Ahead of Print, In-Process & Other Non-Indexed Citations, Daily and Versions(R) (2019 to current);
4. CINAHL (2019 to current);
5. World Health Organization (WHO) COVID‐19 Global Research Database;
6. Centers for Disease Control and Prevention COVID‐19 Research Article Database;
7. International Standard Randomized Controlled Trial Number Registry ([www.isrctn.com](http://www.isrctn.com)).

The neonatal filters were created and tested by the Cochrane Neonatal Information Specialist. The terms for COVID-19 were adapted from the PubMed search used for the Cochrane COVID-19 Study Register (<https://community.cochrane.org/about-covid-19-study-register>).

**CENTRAL via CRS Web:**

**Date searched: 23 July 2020**

Terms:

1 ("2019 nCoV" OR 2019nCoV OR "2019 novel coronavirus" OR “2019-novel CoV” OR ((coronavirus OR "corona virus") AND (Huanan OR Hubei OR Wuhan)) OR "coronavirus-19" OR "coronavirus disease-19" OR "coronavirus disease-2019" OR ((coronavirus or "corona virus") ADJ3 2019) OR "COVID 19" OR COVID19 OR “COVID 2019” OR "nCov 2019" OR ncov19 OR “ncov 19” OR "new coronavirus" OR "new coronaviruses" OR "novel coronavirus" OR "novel coronaviruses" OR "novel coronavirus" OR "novel corona viruses" OR "SARS-CoV2" OR "SARS CoV-2" OR SARSCoV2 OR "SARSCoV-2" OR "SARS-coronavirus-2" OR "SARS-like coronavirus" OR "Severe Acute Respiratory Syndrome Coronavirus-2" OR "severe acute respiratory syndrome coronavirus 2" OR (“spike protein” ADJ3 SARS-CoV-2)) AND CENTRAL:TARGET

2 MESH DESCRIPTOR Infant, Newborn EXPLODE ALL AND CENTRAL:TARGET

3 infant or infants or infant's or "infant s" or infantile or infancy or newborn* or "new born" or "new borns" or "newly born" or neonat* or baby* or babies or premature or prematures or prematurity or preterm or preterms or "pre term" or premies or "low birth weight" or "low birthweight" or VLBW or LBW or ELBW or NICU AND CENTRAL:TARGET

4 MESH DESCRIPTOR Mothers EXPLODE ALL AND CENTRAL:TARGET

5 mother* AND CENTRAL:TARGET

6 maternal AND CENTRAL:TARGET

7 MESH DESCRIPTOR Cesarean Section EXPLODE ALL AND CENTRAL:TARGET

8 (caesarean ADJ3 (section* or deliver*)) AND CENTRAL:TARGET

9 (cesarean ADJ3 (section* or deliver*)) AND CENTRAL:TARGET

10 C-section* AND CENTRAL:TARGET

11 (vaginal* adj3 (birth* or deliver*)) AND CENTRAL:TARGET

12 MESH DESCRIPTOR Delivery, Obstetric EXPLODE ALL AND CENTRAL:TARGET

13 MESH DESCRIPTOR Delivery Rooms EXPLODE ALL AND CENTRAL:TARGET

14 delivery room* AND CENTRAL:TARGET

15 birth suite* AND CENTRAL:TARGET

16 MESH DESCRIPTOR Umbilical Cord EXPLODE ALL AND CENTRAL:TARGET

17 ((umbilicus or umbilical or cord or navel-string) adj2 clamp*) AND CENTRAL:TARGET

18 #2 OR #3 OR #4 OR #5 OR #6 OR #7 OR #8 OR #9 OR #10 OR #11 OR #12 OR #13 OR #14 OR #15 OR #16 OR #17 AND

CENTRAL:TARGET

19 #1 AND #18 AND CENTRAL:TARGET

**MEDLINE via Ovid:**

**Date ranges: 01 January 2019 to 23 July 2020**

Terms:

1. 2019 nCoV.ti,ab.

2. 2019nCoV.ti,ab.

3. 2019 novel coronavirus.ti,ab.

4. 2019-novel CoV.ti,ab.

5. ((coronavirus or "corona virus") and (Huanan or Hubei or Wuhan)).ti,ab.

6. coronavirus-19.ti,ab.

7. coronavirus disease-19.ti,ab.

8. coronavirus disease-2019.ti,ab.

9. ((coronavirus or "corona virus") adj3 "2019").ti,ab.

10. COVID 19.ti,ab.

11. COVID19.ti,ab.

12. COVID 2019.ti,ab.

13. nCov 2019.ti,ab.

14. ncov19.ti,ab.

15. ncov-19.ti,ab.

16. new coronavirus.ti,ab.

17. new coronaviruses.ti,ab.

18. novel coronavirus.ti,ab.

19. novel coronaviruses.ti,ab.

20. novel corona virus.ti,ab.

21. SARS-CoV2.ti,ab.

22. SARS CoV-2.ti,ab.

23. SARSCoV2.ti,ab.

24. SARSCoV-2.ti,ab.

25. SARS-coronavirus-2.ti,ab.

26. SARS-like coronavirus.ti,ab.

27. Severe Acute Respiratory Syndrome Coronavirus-2.ti,ab.

28. COVID-19.rs.

29. COVID-19.rx.

30. COVID-19.kw.

31. COVID-19.kf.

32. coronavirus disease 2019.kw.

33. coronavirus disease 2019.kf.

34. coronavirus disease 19.kw.

35. coronavirus disease 19.kf.

36. corona virus disease 2019.kw.

37. corona virus disease 2019.kf.

38. severe acute respiratory syndrome coronavirus 2.os,ox.

39. severe acute respiratory syndrome coronavirus 2.kw.

40. severe acute respiratory syndrome coronavirus 2.kf.

41. SARS-CoV-2.kw.

42. SARS-CoV-2.kf.

43. COVID-19 drug treatment.ps.

44. COVID-19 drug treatment.px.

45. spike protein, SARS-CoV-2.nm.

46. spike protein, SARS-CoV-2.rn.

47. or/1-46

48. exp infant, newborn/

49. (newborn* or new born or new borns or newly born or baby* or babies or premature or prematurity or preterm or pre term or low

birth weight or low birthweight or VLBW or LBW or infant or infants or 'infant s' or infant's or infantile or infancy or neonat*).ti,ab.

50. exp Mothers/

51. mother*.mp.

52. maternal.mp.

53. exp Cesarean Section/

54. (caesarean adj3 (section* or deliver*)).mp.

55. (cesarean adj3 (section* or deliver*)).mp.

56. C-section*.mp.

57. (vaginal* adj3 (birth* or deliver*)).mp.

58. exp delivery, obstetric/

59. exp Delivery Rooms/

60. delivery room*.mp.

61. birth suite*.mp.

62. exp Umbilical Cord/

63. ((umbilicus or umbilical or cord or navel-string) adj2 clamp*).mp.

64. or/48-63

65. 47 and 64

66. exp Animals/

67. exp Humans/

68. 66 not 67

69. (editorial or comment or letter or newspaper article).pt.

70. 68 or 69

71. 65 not 70

72. limit 71 to yr="2019 -Current"

**Cochrane COVID‐19 Study Register via CRS Web:**

**Date searched: 23 July 2020**

Terms:

1 MESH DESCRIPTOR Infant, Newborn EXPLODE ALL AND INREGISTER

2 infant or infants or infant's or "infant s" or infantile or infancy or newborn* or "new born" or "new borns" or "newly born" or neonat*

or baby* or babies or premature or prematures or prematurity or preterm or preterms or "pre term" or premies or "low birth weight"

or "low birthweight" or VLBW or LBW or ELBW or NICU AND INREGISTER

3 MESH DESCRIPTOR Mothers EXPLODE ALL AND INREGISTER

4 mother* AND INREGISTER

5 maternal AND INREGISTER

6 MESH DESCRIPTOR Cesarean Section EXPLODE ALL AND INREGISTER

7 (caesarean adj3 (section* or deliver*)) AND INREGISTER

8 (cesarean adj3 (section* or deliver*)) AND INREGISTER

9 (C-section or C-sections) AND INREGISTER

10 (vaginal* adj3 (birth* or deliver*)) AND INREGISTER

11 MESH DESCRIPTOR Delivery, Obstetric EXPLODE ALL AND INREGISTER

12 MESH DESCRIPTOR Delivery Rooms EXPLODE ALL AND INREGISTER

13 delivery room* AND INREGISTER

14 birth suite* AND INREGISTER

15 MESH DESCRIPTOR Umbilical Cord EXPLODE ALL AND INREGISTER

16 ((umbilicus or umbilical or cord or navel-string) adj2 clamp*) AND INREGISTER

17 #1 OR #2 OR #3 OR #4 OR #5 OR #6 OR #7 OR #8 OR #9 OR #10 OR #11 OR #12 OR #13 OR #14 OR #15 OR #16

**CINAHL via EBSCOhost:**

**Date ranges: 01 January 2019 to 23 July 2020**

Terms:

("2019 nCoV" OR 2019nCoV OR "2019 novel coronavirus" OR “2019-novel CoV” OR ((coronavirus OR "corona virus") AND

(Huanan OR Hubei OR Wuhan)) OR "coronavirus-19" OR "coronavirus disease-19" OR "coronavirus disease-2019" OR

((coronavirus or "corona virus") N3 2019) OR "COVID 19" OR COVID19 OR “COVID 2019” OR "nCov 2019" OR ncov19 OR “ncov

19” OR "new coronavirus" OR "new coronaviruses" OR "novel coronavirus" OR "novel coronaviruses" OR "novel corona virus" OR

"novel corona viruses" OR "SARS-CoV2" OR "SARS CoV-2" OR SARSCoV2 OR "SARSCoV-2" OR "SARS-coronavirus-2" OR

"SARS-like coronavirus" OR "Severe Acute Respiratory Syndrome Coronavirus-2" OR "severe acute respiratory syndrome

coronavirus 2" OR (“spike protein” N3 SARS-CoV-2)) AND

(infant or infants or infant’s or infantile or infancy or newborn* or "new born" or "new borns" or "newly born" or neonat* or baby* or

babies or premature or prematures or prematurity or preterm or preterms or "pre term" or premies or "low birth weight" or "low

birthweight" or VLBW or LBW or mother* or maternal or (caesarean N3 (section* or deliver*)) or (cesarean N3 (section* or deliver*))

or C-section* or (vaginal* N3 (birth* or deliver*)) or delivery room* or birth suite* or ((umbilicus or umbilical or cord or navel-string)

N2 clamp*))

Limiters - Published Date: 20190101-20201231

**World Health Organization (WHO) COVID-19 Global literature on**

**coronavirus disease:**

(<https://search.bvsalud.org/global-literature-on-novel-coronavirus-2019-ncov>)

**Date searched: 23 July 2020**

Terms:

infant or infants or infant’s or infantile or infancy or newborn* or "new born" or "new borns" or "newly born" or neonat* or baby* or

babies or prematures or prematurity or preterm or preterms or "pre term" or premies or "low birth weight" or "low birthweight" or

VLBW or LBW or mother* or maternal or caesarean or cesarean or “vaginal birth” or “vaginal delivery” or “vaginal deliveries” or

“delivery room” or “delivery rooms” or “birth suite” or “birth suites” or “umbilical cord clamping” or "cord clamping"

Centers for Disease Control and Prevention COVID-19 Research

Articles Database via EndNote database:

(<https://www.cdc.gov/library/researchguides/2019novelcoronavirus/researcharticles.html>)

Date searched: 23 July 2020

Terms:

infant*

Baby

babies

neonat*

Newborns

Preterm

mother*

Maternal

caesarean

cesarean

C-section

C-sections

vaginal birth

vaginal delivery

vaginal deliveries

delivery room

delivery rooms

birth suite

birth suites

Umbilical cord clamping

cord clamping

**ISRCTN:**

**Date searched: 24 July 2020**

Terms:

covid-19 AND ( Participant age range: Neonate )

covid-19 AND infant*

covid-19 AND newborns

covid-19 AND neonat*

covid-19 AND preterm*

covid-19 AND "delivery room*"

covid-19 AND vaginal delivery

covid-19 AND vaginal birth

covid-19 AND caesarean

covid-19 AND cesarean

covid-19 AND mother*

covid-19 AND maternal

covid-19 AND cord clamping

**Updated Search Strategy**

Comparison of the initial search strategy and an updated search strategy using PubMed and Google Scholar showed that 96.5% of total searched articles were captured using the PubMed and Google updated search approach. Two author reviewers conducted this updated search strategy (LPB, JMH). Updates through PubMed and Google Scholar were subsequently conducted July 23, 2020 until end date December 31, 2021.

**PubMed key word search strategy**: (*year published) (covid-19 OR SARS-cov-2 OR coronavirus) AND (neonatal OR newborn OR neonates OR infants OR pregnancy OR (vertical transmission))*

**Google Scholar key word search strategy**: ((covid-19 OR SARS-cov-2 OR coronavirus) AND (neonatal OR newborn OR neonates OR infants OR pregnancy OR (vertical transmission))) AND (("2021"[Date - Publication] : "2021"[Date - Publication])) NOT (Cohort OR Cohorts) NOT (Commentary OR Commentaries) NOT (Editorial OR Editorials) NOT (analysis OR meta-analysis) NOT (Review OR Reviews) NOT (Vaccine OR Vaccines OR Immunization) NOT (Test OR testing)

**Appendix 3: Detailed Description of Delivery Management Interventions.**

**Category A: Physical environment - aerosolization and droplet management**

1. Presence of negative pressure in delivery room

Negative pressure refers to the pressure differential creating air flow into a patient room whenever the door is opened. This is recommended to prevent aerosolized particles generated in the patient room from contaminating areas outside the patient environment ^1^. Given the concern that SARS-CoV-2 may transmit through aerosol and contact surfaces, negative pressure delivery rooms have been recommended for any laboring mother who is SARS-CoV-2-positive ^2, 3^. As of January 2022, it remains unclear whether a negative pressure room reduces the risk of early neonatal infection.

2. Location for neonatal resuscitation

There is currently limited knowledge on the optimal location for neonatal resuscitation, especially when advanced resuscitation requiring a larger resuscitation team is anticipated due to a preterm birth or complicated perinatal course. Immediate separation from an infected mother could potentially minimize the risk of exposure to the neonate. However, providing infant resuscitation at a mother's bedside has been shown to have beneficial effects on infant transition because of the ability to do immediate skin-to-skin contact and delayed cord clamping ^4 usability and acceptability of a mobile trolley^. Mother-infant bonding is also promoted by having the baby nearby during the transition period. In situations where resuscitation is required, family presence has been shown to improve family understanding during cardiopulmonary resuscitation ^5^. Separation by room or distance decreases family-centered care and may increase anxiety, decrease bonding, and create distrust of the medical team.

3. Maternal masking during labor

Maternal masking has been suggested as a method for reducing the aerosolized respiratory droplets that may be formed especially during the second and third stages of labor ^6^. Maintaining this mask is difficult especially in situations where a mother may require additional oxygen for the progression of labor. The risks and benefits of such practice as it pertains to early neonatal infection remains hypothetical.

**Category B: Delivery specific interventions - minimization of contact during delivery with maternal fluids**

1. Mode of delivery

Caesarean delivery has been suggested as a mode to minimize or avoid neonatal exposure to maternal vaginal secretions, urine, or stool where SARS-CoV-2 viral load is presumably higher based on extrapolations from HIV prevention measures ^7^. If high viral load is present, vertical transmission risk might theoretically decrease with caesarean section done after shorter duration of ruptured membranes. This potential benefit may be negligible, especially if low/undetectable viremia was associated with lower risk of vertical transmission, as seen with HIV. Importantly, caesarean delivery may increase the risk of droplet or airborne spread in the case of endotracheal intubation for maternal anesthesia, high-flow oxygen or electrocautery of the surgical wound ^8^.

Caesarean delivery for infection prevention will also increase the medical morbidity to the mother, as well as the cost and complexity of this and future pregnancies ^9^. It is also well known that caesarean birth is associated with high risk for neonatal respiratory complications secondary to the delayed transition of fetal to newborn adaptation ^10^.

2. Timing of cord clamping

Delayed cord clamping, as defined by 30 seconds or more, has been demonstrated to have beneficial effects for both preterm and term infants ^11-14^ and is currently recommended for all infants who do not need resuscitation at birth ^15^. However, it may have the unintended effect of increasing transplacental viral passage and prolonged infant exposure to maternal secretions because of positioning the neonate within close proximity of the mother.

Conversely, early cord clamping as defined by less than 30 seconds from delivery of infant to cord clamping, may reduce viral transmission and avoid prolonged exposure of infant to infected maternal secretions or stool. With delayed cord clamping being the current standard of care (in most countries and hospitals), early cord clamping may potentially deprive the preterm neonate of the benefits of risk reduction in necrotizing enterocolitis (NEC), intraventricular hemorrhage (IVH), or even death ^13^.

**Category C: Infant care practices - minimizing infant skin contact**

1. Skin-to-skin contact

Immediate mother-to-infant skin-to-skin contact after delivery has been shown to provide benefits to the mother and infant dyad. For the postpartum mother, there is evidence that it can accelerate the third stage of labor and in turn prevent postpartum hemorrhage ^16^. Skin-to-skin contact right after birth can also improve newborn vital signs including temperature stability, promote bonding, and improve breastfeeding success ^17^. With concerns of infectious risk from a SARS-CoV-2 infected mother to her infant, several countries have recommended against skin-to-skin care in order to minimize direct neonatal exposure to droplets ^18^.

2. Decontamination of neonate after delivery

Cleaning the infant immediately after birth may theoretically reduce the risk of transmission especially when mother is viremic and the exposed infant has direct contact with maternal bodily fluids like blood and amniotic fluid infected with SARS-CoV-2. However, immediate cleaning will decrease the availability of vernix which has many beneficial effects for newborn infant skin health including a barrier to prevent water loss, temperature regulation, innate immunity and even promotion of healthy intestinal development ^19, 20^. In addition, cleaning extremely low gestational age infants might increase the risk of hypothermia and skin breakdown, especially when antiseptic solutions are used ^21^.

**Appendix 4. Clinical Status Classifications for SARS-CoV-2 Positive** **Patients**

**Pediatric Patients**: Dong, et al. describes the clinical characteristics of more than 2000 pediatric patients and divides their clinical status into asymptomatic, mild, moderate, severe and critical ^22^:

1. Asymptomatic: without clinical signs and symptoms and chest imaging is normal.
2. Mild: symptoms of acute upper respiratory tract infection, including fever, fatigue, myalgia, cough, sore throat, runny nose, and sneezing. Physical examination shows congestion of the pharynx without auscultatory abnormalities. Some may present with digestive symptoms such as nausea, vomiting, abdominal pain, and diarrhea.
3. Moderate: diagnosed with pneumonia, with fever and cough, wheezing, and upper respiratory congestion without hypoxemia. Some cases may be asymptomatic but will have chest CT or X-ray showing subclinical lung lesions.
4. Severe: hypoxia with other previously described symptoms with or without imaging findings.
5. Critical: Multi-organ dysfunction with respiratory failure, shock, encephalopathy, myocardial impairment, coagulation dysfunction or kidney injury.

**Maternal Patients**: Severity of maternal COVID-19 symptoms have been described in levels from asymptomatic to critical based on the Berlin Definition for Acute Respiratory Distress Syndrome ^23^.

1. Asymptomatic;
2. Mild: any signs and symptoms (e.g. fever, cough, sore throat, malaise, headache, muscle pain) without shortness of breath, dyspnea, or abnormal chest imaging);
3. Moderate: lower respiratory disease by clinical assessment or imaging and a saturation of oxygen > 93% on room air;
4. Severe: respiratory frequency > 30 breaths per minute, SaO_2_ ≤ 93% on room air, ratio of arterial partial pressure of oxygen to fraction of inspired oxygen (PaO_2_ /FiO_2_) < 300, or lung infiltrates > 50%;
5. Critical: respiratory failure, septic shock, and/or multiple organ dysfunction.

**Appendix 5: List of articles included by year for final analysis.**

**2020 Articles Included: Total 93**

8. Alzamora MC, Paredes T, Caceres D, Webb CM, Valdez LM, La Rosa M. Severe COVID-19 during Pregnancy and Possible Vertical Transmission. *Am J Perinatol*. 2020.

22. Chhabra A, Rao TN, Kumar M, Singh Y, Subramaniam R. Anaesthetic management of a COVID-19 parturient for caesarean section - Case report and lessons learnt. *Indian J Anaesth*. 2020;64(Suppl 2):S141-S143.

23. Correia CR, Marcal M, Vieira F, Santos E, Novais C, Maria AT, et al. Congenital SARS-CoV-2 Infection in a Neonate With Severe Acute Respiratory Syndrome. *Pediatr Infect Dis J*. 2020;39(12):e439-e443.

24. De Socio GV, Malincarne L, Arena S, Troiani S, Benedetti S, Camilloni B, et al. Delivery in Asymptomatic Italian Woman with SARS-CoV-2 Infection. *Mediterr J Hematol Infect Dis*. 2020;12(1):e2020033.

25. Díaz CA, Maestro ML, Pumarega MTM, Antón BF, Alonso CP. First case of neonatal infection due to COVID 19 in Spain. *An Pediatr (Engl Ed)*. 2020.

48. Lee DH, Lee J, Kim E, Woo K, Park HY, An J. Emergency cesarean section on severe acute respiratory syndrome coronavirus 2 (SARS- CoV-2) confirmed patient. *Korean J Anesthesiol*. 2020.

49. Lee EK, Kim WD, Lee DW, Lee SA. Management of the first newborn delivered by a mother with COVID-19 in South Korea. *Clin Exp Pediatr*. 2020.

61. Nawsherwan, Khan S, Nabi G, Fan C, Wang S. Impact of COVID-19 Pneumonia on Neonatal Birth Outcomes. *Indian J Pediatr*. 2020.

74. Sinelli MT, Paterlini G, Citterio M, Di Marco A, Fedeli T, Ventura ML. Early Neonatal SARS-CoV-2 Infection Manifesting With Hypoxemia Requiring Respiratory Support. *Pediatrics*. 2020.

83. Wang X, Zhou Z, Zhang J, Zhu F, Tang Y, Shen X. A case of 2019 Novel Coronavirus in a pregnant woman with preterm delivery. *Clin Infect Dis*. 2020.

**2021 Articles Included: Total 24**

10. Komiazyk M, Aptowicz A, Książek I, Sitkiewicz I, Baraniak A. An asymptomatic carriage of severe acute respiratory syndrome coronavirus 2 by a pregnant woman and her newborn. *Polish Archives of Internal Medicine*. 2021.

**Appendix 6: Subgroup analysis of maternal illness severity and gestational age.** This lists frequencies of primary outcomes for cases describing maternal illness severity and infant gestational age described in large categories <34 weeks gestational age (GA) or ≥34 weeks GA.

| **Subgroup analysis** | **n/N** | **Early Neonatal Infection, percentage [95% confidence interval]** | **n/N** | **Neonatal Death, percentage [95% confidence interval]** |
| --- | --- | --- | --- | --- |
| **Maternal Illness Severity:**  **n/total (%)** |  |  |  |  |
| Asymptomatic | 5/29 | 30.66 [ 6.87; 54.45] | 0/29 | 0.00 [ 0.00; 8.72] |
| Mild | 28/84 | 42.39 [28.53; 56.24] | 2/84 | 1.50 [ 0.00; 7.30] |
| Moderate | 6/68 | 8.74 [ 0.00; 17.50] | 1/67 | 0.60 [ 0.00; 6.94] |
| Severe | 8/24 | 33.33 [12.60; 54.07] | 1/24 | 4.15 [ 0.00; 16.38] |
| Critical | 7/33 | 22.82 [ 7.36; 38.29] | 1/32 | 3.04 [ 0.00; 13.50] |
| **Gestational Age:**  **n/total (%)** |  |  |  |  |
| Moderate or very preterm (<34 weeks GA) | 19/53 | 33.70 [18.92; 48.48] | 4/52 | 6.82 [ 0; 15.10] |
| Term/late preterm (≥34 weeks GA) | 33/183 | 27.41 [18.40; 36.42] | 1/182 | 0.22 [ 0; 3.94] |
